# Quantifying the intangible: Evidence from Nigeria on the impact of supervision, autonomy, and management practices on PHC performance in the context of Direct Facility Financing

**DOI:** 10.1101/2024.08.15.24312076

**Authors:** Brittany Hagedorn, Benjamin Loevinsohn, Oluwole Odutolu

## Abstract

**Key messages:**

- Secondary analysis of data from the Nigeria States Health Investment Project showed improvements in facility autonomy, budget control, and management practices in experimental (PBF and DFF) districts much more so than in control districts.
- Supportive supervision, including use of a quantified checklist, contributed to improvements in facility readiness to provide services and clinical productivity.
- Primary healthcare facilities with better management practices had improved service readiness and offered more services than those with worse management practices.

Previous studies have shown that facility autonomy, especially control over budget allocation, can have a modest positive effect on performance, but the findings depend on the context. Similarly, management practices are often cited as important contributors to facility performance, but the evidence is limited and usually qualitative. Data from the large-scale randomized evaluation of the Nigeria States Health Investment Project (NSHIP) offers an opportunity to quantitatively examine these relationships in the context of a lower middle-income country. We utilize non-parametric statistics to test for difference in means and apply regression analysis to test the hypothesis that autonomy and management affected facility performance. Our results show that facilities with greater autonomy, more budget control, and better management practices generally outperform their peers on a range of facility readiness and service delivery measures. For example, regression results found that facilities with high autonomy held on average 2.1 more outreach sessions per month than those without, and facilities with an annual business plan offered 1.8 additional outreach services. Supervision practices, such as more frequent visits and use of a quantitative checklist, are associated with 26% higher productivity and up to a 28.6% increase in equipment availability (percentage points), respectively. We conduct sensitivity analyses on our variable selection and use a random forest approach to validate that results are robust to changes in the model structure. We conclude that facility-level autonomy and especially budget control can improve primary healthcare facility readiness and service availability, even in resource-constrained contexts, Further, this can be achieved through good management practices that are reinforced through supportive supervision and routine performance monitoring to maximize the gains that result from incremental financing. This shows that these policies and practices can be critical contributors to efficiently achieving the goals of universal healthcare policies in the context of limited resources.

## Introduction

There is an ongoing discussion about the value of facility-level autonomy over budgets and allocation decisions in improving primary healthcare (PHC) (Collins and Green 1994). In theory, local management should be helpful in ensuring that PHC systems meet local needs in culturally-sensitive ways and respond flexibly and rapidly to contextual challenges. This adaptability is thought to stem from community engagement, enabling people who are under-served to access decision-makers, and creating opportunities for direct participation in decision making, such as through facility health committees (WHO 2022).

There has been a growing movement to shift decision-making closer to healthcare users (WHO 2022). There are reasons to expect that local autonomy and management would impact PHC performance, for example that district control over drug procurement policies would guide facility product stocking behavior and affect stock-out levels.

On aggregate, financial autonomy at the facility level has been found to be necessary but not sufficient to guarantee good outcomes (DeWalque et al. 2022, Mabuchi et al. 2022). A comprehensive World Bank review of performance-based financing (PBF) initiatives found that decentralized facility financing (DFF) and autonomy improved effective coverage rates when paired with strong supervision and community oversight (Gage and Bauhoff 2021).

This is supported by case studies such as in Kenya, where facilities that retain and manage their revenues report better service readiness (Bonfiace et al. 2023) and in Indonesia, where granting facilities the ability to receive, retain, and use funds for infrastructure and operational investments has been linked to performance improvements (Rawung and Sholihin 2017). Further, management behaviors have been shown to impact facility readiness to deliver family planning services in Mozambique (Pope et al. 2017).

However, positive outcomes depend on the context (Collins and Green 1994, Das et al. 2016). For example, in Tanzania, health facility management committees have a positive but inconsistent impact on health system performance et al. 2022). In Kenya, it has been important to have supporting functions like public financial management in place (Barawa et al. 2022). In Ghana, management skills are associated with better process and patient experience outcomes (Macarayan et al. 2019).In India strong management, but not autonomy, was associated with positive performance (Desai et al. 2022).

We apply WHO’s definition of financial autonomy as ensuring that a facility can receive funds, manage them, and make operational spending decisions. This includes the right to procure inputs such as supplies and human resources (WHO 2022). Autonomy should not be confused with decentralization, service purchasing or insurance, or provider payment systems.

In the cluster randomized evaluation of the Nigeria State Health Investment Project (NSHIP) which has been described in detail elsewhere (Khanna et al. 2021), management practices, supervision quality, and facility financial autonomy were all credited as important to the program’s success compared to the control group. In the midpoint qualitative review of high performing facilities, respondents indicated that supervision had improved in both frequency and quality, managers of high-performing PHCs investigated and resolved performance issues, and performance bonuses were used to incentivize improvements to utilization and service quality in the PBF arm (World Bank 2015). The importance of autonomy, strengthened supervision, and improved management practices was highlighted by the similarity in results achieved by DFF and PBF facilities, given that both study arms had management- and autonomy-related interventions (Kandpal et al. 2018).

These findings about autonomy, supervision, and management are based on qualitative reporting and expert views. In this study, we examine the effects of these factors quantitatively using data from NSHIP’s facility surveys. If they are found to be significant predictors of PHC readiness in Nigeria, this indicates that they should be included in future models of PHC systems. Additionally, quantifying their effect sizes enables more direct comparison with other investments in health that have more easily attributable impacts (e.g. provision of in-kind supplies). Insights gained from this analysis will enhance the existing literature by providing a nuanced depth of understanding on unstudied components of PHC systems. Additionally, quantified estimates of the effect sizes of funding level, financial incentives, budgetary autonomy, and state-level contextual factors can inform allocation of limited resources and ultimately create stronger health systems.

## Materials and methods

### Data

We used the NSHIP independent health facility surveys conducted in 2014 and 2017 with the latter considered the ‘endline’ assessment conducted after three years of implementation of PBF and DFF (Kandpal 2021). The facility survey form known as “HF1” was a comprehensive assessment of infrastructure, supplies, human resources, management practices, etc.

We used the HF1 survey section six, titled “autonomy” (see Supplement 1 for the specific questions), which asked the health facility manager nine questions regarding how work was organized and decisions made. Respondents answered whether they felt nine statements were true ‘most of the time’, ‘more than half of the time’, ‘less than half of the time’, ‘rarely’ or ‘never’. For example, one of the statements was: “I have enough authority to obtain the resources I need (drugs, supplies, funding) to meet the needs of my facility.”

We transform these responses into two aggregate measures and two individual question binaries. This structure was based on a combination of expert opinion and the qualitative statements in the NSHIP impact and evaluation report and previous literature, which shows that quality of supervision and budget autonomy are important for PHC facility outcomes (World Bank 2015, Kandpal et al. 2018).

The four variables were defined as:

- Aggregate high autonomy = The percent of questions (of nine) that were answered with “more than half” or “most of the time”.
- Aggregate low autonomy = The percent of questions (of nine) that were answered with “rarely” or “never”.
- Budget control = Based on question 6.01, “I am able to allocate my facility budget according to how it is needed. There is enough flexibility in my budget.”. A value of one if the respondent indicated “more than half” or “most of the time”; otherwise zero.
- Supervision quality = Based on question 6.09, “The HMB/LGA/PHC Department Team provides adequate feedback to me about my job and the performance of my facility.” A value of one if the respondent indicated “more than half” or “most of the time”; otherwise zero.

We also considered questions from section 2 of the facility survey relating to management practices, including:

- The presence, composition, and frequency of meetings of the facility’s PBF/DFF committee.
- The presence, composition, frequency of meetings, and quality of meeting records of the facility’s management team.
- The existence of a facility budget and business plan, and whether there was a prioritization of health activities included them.
- The existence of written job descriptions for facility staff.
- Frequency of supervision for the facility and staff.
- Whether the facility tracks its performance for any set of indicators.

We checked for collinearity amongst the survey questions using a Pearson correlations test and removed variables from consideration as appropriate.

We use the term ‘supervised autonomy’ as shorthand for the complex decision-making and management processes captured by these nine questions. In fact, they incorporate a range of structural mechanisms that includes budgeting processes, supportive supervision, written policies and procedures, resource mobilization, and operational flexibility. We use the shorthand for ease of reporting results and discussing implications, but do not intend to ignore or over-simplify the reality of management practices.

### Impact of external supervision

Supervision visits were conducted in both PBF and DFF facilities using a structured, quantitative checklist. In PBF facilities, the quarterly results from the checklist went into calculating performance payments. Theoretically, the increases in structural quality that were reported in the impact evaluation (Kandpal et al. 2018) may have been due primarily to this supervision process and not in response to facility needs. We examine how local facility managers were making decisions on how to spend their resources – solely using the checklist or not – by looking at changes in equipment availability (Questions 15.02 A-Y) by item. We use a t-test to compare changes in the distribution means between baseline and endline for each item and report the aggregate changes for those items on and not on the supervision checklist.

### Impact of supervision on productivity

To further examine the role of supervision, we also explored the role that supervision frequency had on individual clinical productivity. In theory, a well-managed clinical staff (i.e. nurses, midwives, and community health workers) would be able to perform better in their roles, thus potentially increasing their productivity. This is measured in the staff survey by asking each individual how many patients they treated on their previous full day of work. We examine the bivariate relationship between reported supervision frequency and productivity, grouping the frequency into categories approximating yearly, quarterly, monthly, or more than monthly.

### Impact of budget allocation control

Both PBF and DFF facilities were provided with facility-level bank accounts and considerable autonomy over how funds were spent. To examine the impact of budget control on outcomes, we compare the change in equipment availability between 2014 and 2017 for facilities with differing levels of supervised autonomy and specifically budget control. Level of supervised autonomy was categorized as follows: very high = all question responses “more than half the time” or “most of the time”, low = more than half of question responses “rarely” or “never”, moderate = between the two extremes.

### Regression approach

We used a linear regression approach to test whether the four supervised autonomy-related independent variables were significantly associated with changes in facility outcomes, such as structural quality, after controlling for confounders including state, study arm, and facility revenues. Study arm was defined categorically as: control, DFF, or PBF. Facilities revenues were reported in the survey for 2016 and we normalize them per capita based on the facility-reported catchment population.

The outcome variables were aggregated from the facility survey and include all sections pertaining to structural quality and service offerings, such as functional equipment availability, products in stock, hours of antenatal care per week, and the number of services offered via outreach. Detailed outcome descriptions are listed in Supplement 2. We also used the total outpatient volume as an independent outcome, representing overall service utilization.

Including potential interaction terms, the final regression is of the form:

Outcome = autonomy variables + management practices + control variables + interactions, where:

- Autonomy variables: budget control + supervision quality + high autonomy + low autonomy
- Management practices: questions from survey section 2 (see data section above)
- Control variables: state + study arm + revenue
- Interactions: autonomy*study arm + autonomy*state + revenue*budget control + autonomy*budget control

If an independent variable was significant in a greater proportion of outcome regressions than random chance would imply, we concluded that it is a predictor of facility readiness. We removed any terms that were not significant above a random chance of five percent, re-ran the regression models, and present those updated results.

Regressions were run in R Studio version 2023.03.0 Build 386, using the function stepAIC from the MASS package for automated backward selection of the independent variables (R Core Team 2023, Ripley et al. 2023). The final data sample included 980 PHC facilities across six states, including both control and intervention facilities.

### Sensitivity analysis

There were three potential concerns that could have biased these results and we conducted sensitivity tests on each of them.

First was the possibility that the estimated impact of autonomy was in fact due to other factors associated with the study, such as the PBF incentives or additional revenues. To test this, we ran the regressions without any autonomy variables.

Second was the possibility that the way we designed the independent variables for autonomy influenced the results. This is mitigated somewhat by the fact that aggregation could obscure some important heterogeneity and is most likely to reduce our ability to detect a relationship, thus making our estimates a conservative estimate of effect size. However, we still wanted to test this theory, so we also ran the regressions with the independent survey responses (normalized on a scale of 0-1) instead of the four variables as previously defined.

We used analysis of variance (ANOVA) to test both alternative regression structures compared to the approach used in the core analysis.

Third was the possibility that in fact the relationship between the independent and dependent variables is not linear (or logged, in the case of revenues) and that our choice to use regressions was inadequate. To test this, we opted to replicate the analysis using a random forest approach, which is a method with different assumptions and has the benefit of allowing multiple threshold values for which an independent variable is meaningful and does not require linearity.

## Results

### Impact of external supervision

In the control arm, the percentage of facilities with functional equipment did not change meaningfully across the board, with eleven of the twenty-five equipment items (44%) decreasing in availability between baseline and endline. In the intervention arm, all but one item (incubators) increased in availability (Figure 1).

**Figure 1.**
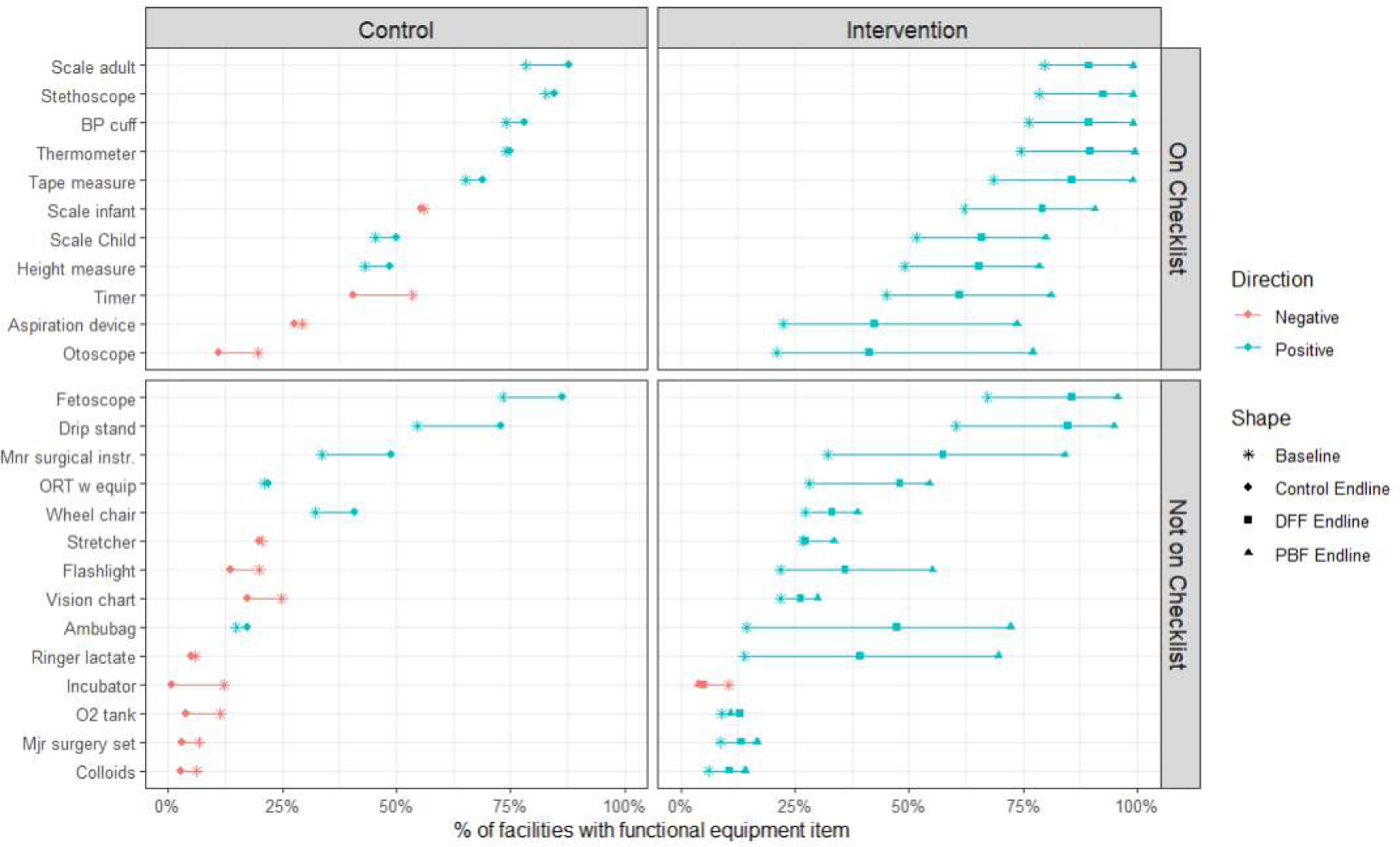
Percentage of facilities with a functional equipment item, by time point, study arm, and presence on supervision checklist. Calculated based on observed equipment in facility, survey questions 15.02 A-Y. Baseline data is from the independent facility survey conducted in 2014. Endline data is from the independent facility survey conducted in 2017. Checklist grouping indicates whether a piece of equipment was present on the quarterly supervision checklist that was used to determine incentive payments. Control facilities were in Benue, Ogun, and Taraba. Intervention facilities were in Adamawa, Nasarawa, and Ondo. DFF = direct facility financing. PBF = performance-based financing. BP = blood pressure. Mjr = major. O2 = oxygen. ORT = oral rehydration therapy. Mnr = minor.

The median increase in equipment availability varied by study arm. In the PBF arm, the median increase in availability was 28.6% for items on the checklist and 19.0% for those not. In the DFF arm, availability increased by 16.0% and 10.0% (percentage points) for items on the checklist or not, respectively. In the control arm, this was 1.9% and −0.1%, respectively.

In the intervention arm facilities, all of the eleven equipment items that were on the supervision checklist had a statistically significant increase in availability in both DFF and PBF study arms (p-value <0.001). Of items not on the checklist, eight of fourteen items (57%) increased in availability in DFF facilities and eleven of fourteen items (79%) increased in PBF facilities.

### Impact of supervision on productivity

We observe that there does appear to be a relationship between supervision frequency and clinical productivity, although we cannot confirm causality. Specifically, there was an average of 27% higher level of reported productivity in nurses and midwives, and 26% higher productivity in community health extension workers, when comparing those with more or less than monthly supervision visits. There is a statistical difference in means (Wilcox test) for CHEW (p=0.01) but not for nurses and midwives (p=0.07). This was true both at baseline and in the endline surveys. The summary statistics are shown in Figure 2.

**Figure 2.**
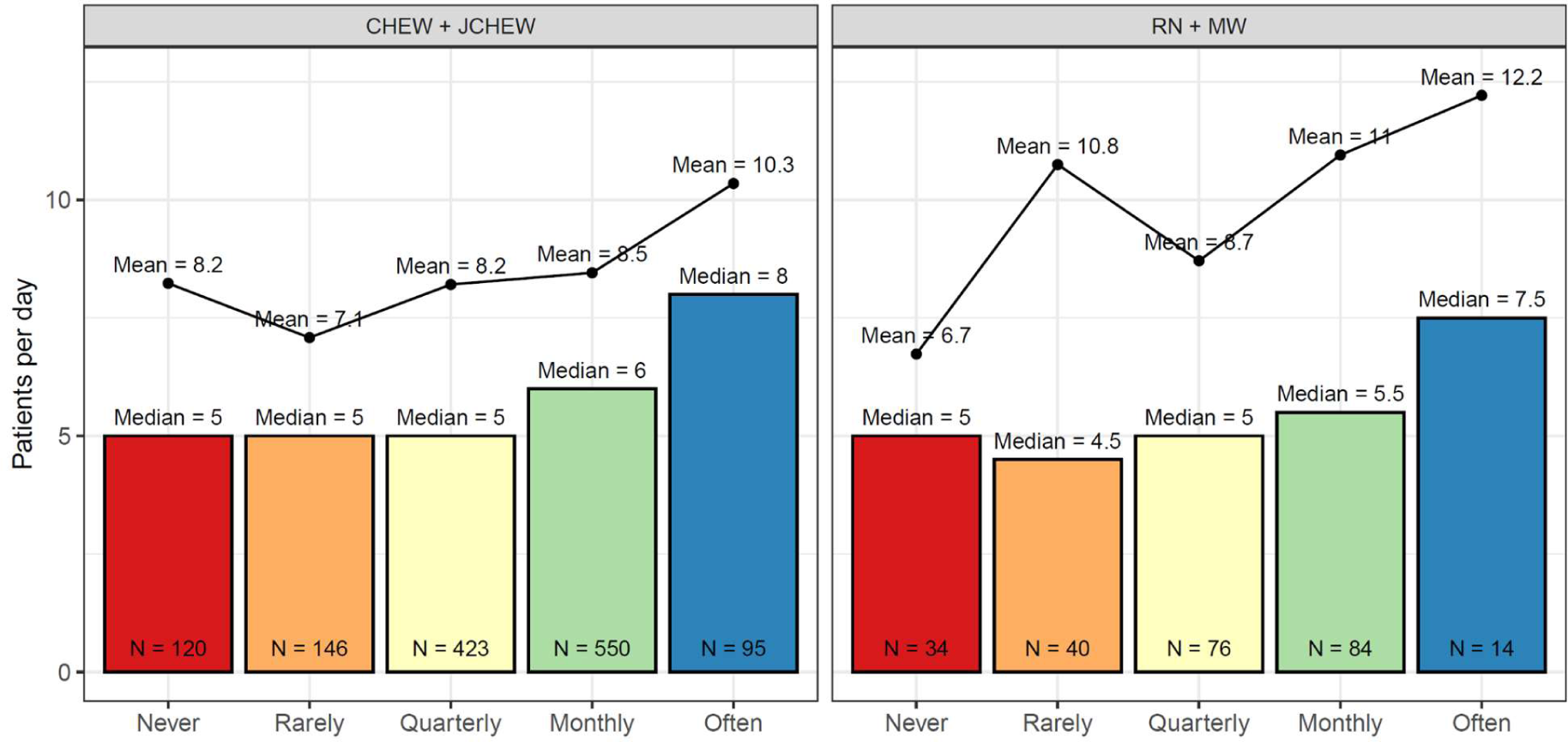
Relationship between supervision frequency and clinical staff productivity, 2017. Mean and median values are higher for those with supervision more often than monthly. Productivity was reported by individual staff members based on the number of patients seen in their previous day of work, excluding staff who had not worked at least one day in the last 30 days. Supervision frequency was also reported by the staff member. N = sample size. CHEW = community health extension worker. JCHEW = junior community health extension worker. RN = nurse. MW = midwife.

### Impact of budget allocation control

In intervention facilities (DFF and PBF combined), there was no statistically significant difference in the mean equipment availability between facilities with high and moderate supervised autonomy responses. However, facilities with low autonomy (those reporting ‘rarely’ or ‘never’ on more than half of the nine survey questions), were less well-equipped than their peers at a p-value of <0.01.

Regardless of the overall level of autonomy, budget control is associated with larger increases in equipment availability. In facilities with budget control, the high and moderate autonomy categories, the median increased from nine and ten at baseline to fifteen at endline, an increase of five items (50-55%). In facilities without budget control, the median in moderate autonomy facilities increased from ten to twelve items (20%), while low autonomy facilities decreased from eight to five (−38%). (Figure 3) The increase in performance from baseline to endline within each group was statistically significant for the very high and moderate autonomy sub-groups (Wilcox test, p<0.001). Among facilities with moderate overall autonomy, there was not a significant difference between facilities with or without budget control at baseline (p=0.40), but there was at endline (p<0.001).

**Figure 3.**
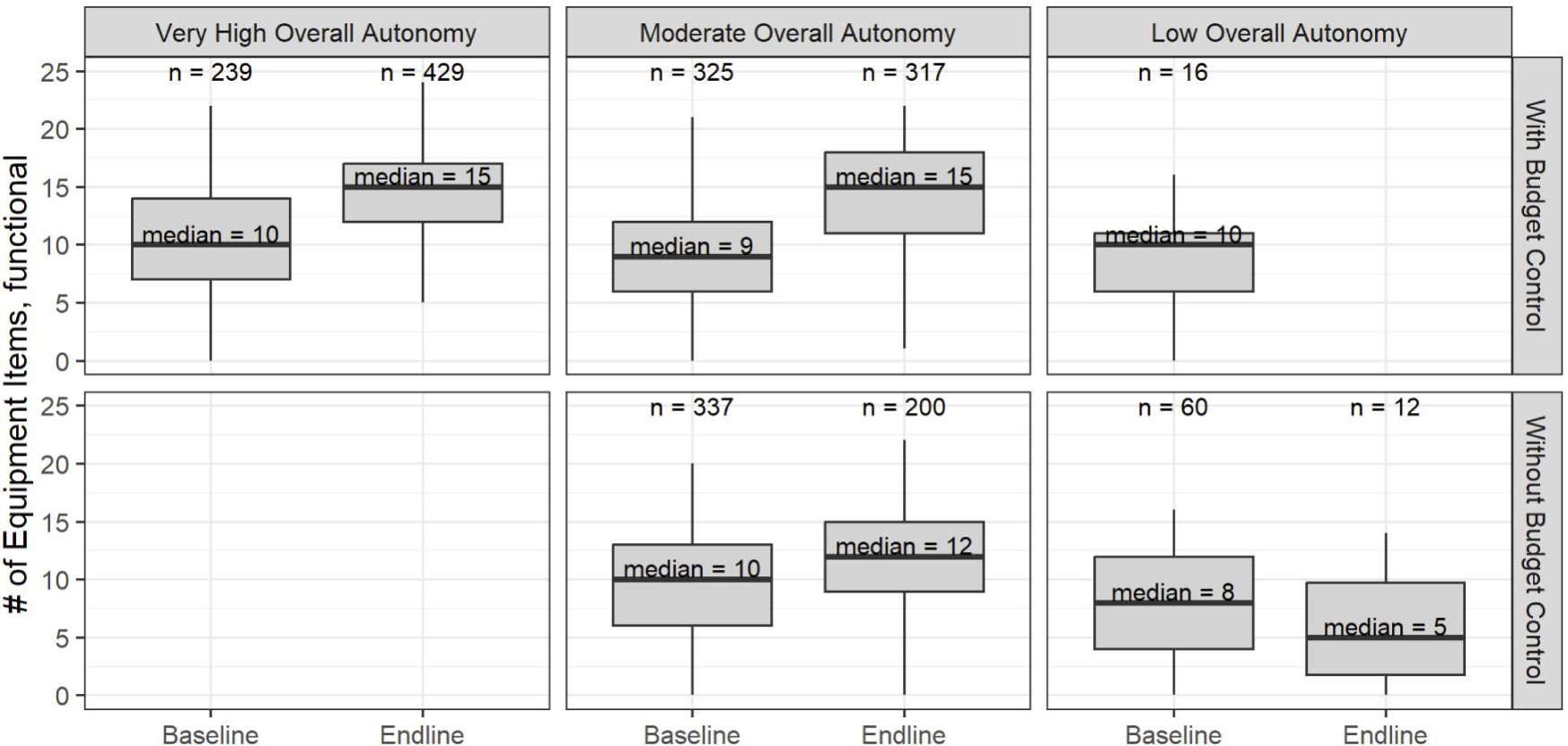
Impact of budget autonomy in combination with supervised autonomy on equipment availability. Number of equipment items calculated based on the potential list of 25 from the facility survey questions 15.02 A-Y. n = sample size. Budget control classification based on question 6.01. Autonomy classification based on questions 6.01-6.09. Boxplot central line is the distribution median, box upper and lower edges depict 25th and 75th percentiles, and lines extend to the 5th and 95th percentiles. The plot only shows subsets with sample size of five or more facilities with complete data, which reduced the total sample by less than one percent.

Comparisons of control arm facilities follow similar patterns. Additionally, Wilcox tests of difference in means showed that facilities with moderate to high autonomy had significant gains in intervention facilities but not in control facilities (details in Supplement 4).

## Regression Results

Of the 980 level 1 PHC facilities included in the study, 742 reported population and revenues; of those, 729 facilities had complete data on autonomy and management practices and were included in the regressions (244 DFF, 361 PBF, 124 control). A review of overall characteristics of the facilities that were excluded did not indicate a non-random selection bias.

After examining the results, several potential independent variables were removed because they were not significant with a frequency above random chance. These included the interaction terms between autonomy and state, revenue and budget control, and overall autonomy and budget control, as well as the number of members on the PBF/DFF committee, specific membership in the PBF/DFF and health facility management committees, frequency of staff meetings, and initiatives undertaken by the health facility committee in the last twelve months. We reran the regressions without these variables included, which are the results presented below.

For total outpatient visits, both regressions with and without adjustment for supervised autonomy had adjusted R squared values of 0.50 and the ANOVA comparison between the two regressions was insignificant. In the autonomy regression, budget control and the percent of questions answered with ‘high autonomy’ were statistically significant with p=0.017 and p=0.003 respectively. However, this did not improve the overall fit of the model.

State is the most frequently significant regression variable, followed by revenues per capita and study arm, both of which are significant in approximately half of regressions at the p<0.05 level. In addition, the interaction terms between revenue and study arm are also significant in 7 (DFF) and 5 (PBF) regressions.

The variables of interest for this study, pertaining to supervised autonomy and management practices, are also found to be significant at a rate above random chance. Autonomy variables were significant in about one third of regressions at p < 0.05, slightly less than study arm and revenues. Five management practices are significant in at least half of regressions and another five are significant in at least a quarter (Figure 4). The most frequently significant predictors were related to the existence of the PBF/DFF committee and health facility management committee, the frequency of meetings and external supervision, and having an annual business plan. Note that not all variables that were evaluated were significant. For example, the presence of a member of the ward development council and a school headmaster on the health facility management committee were significant, but health facility staff were not. Coefficients and p-values for these regressions are detailed in Supplement 3.

**Figure 4.**
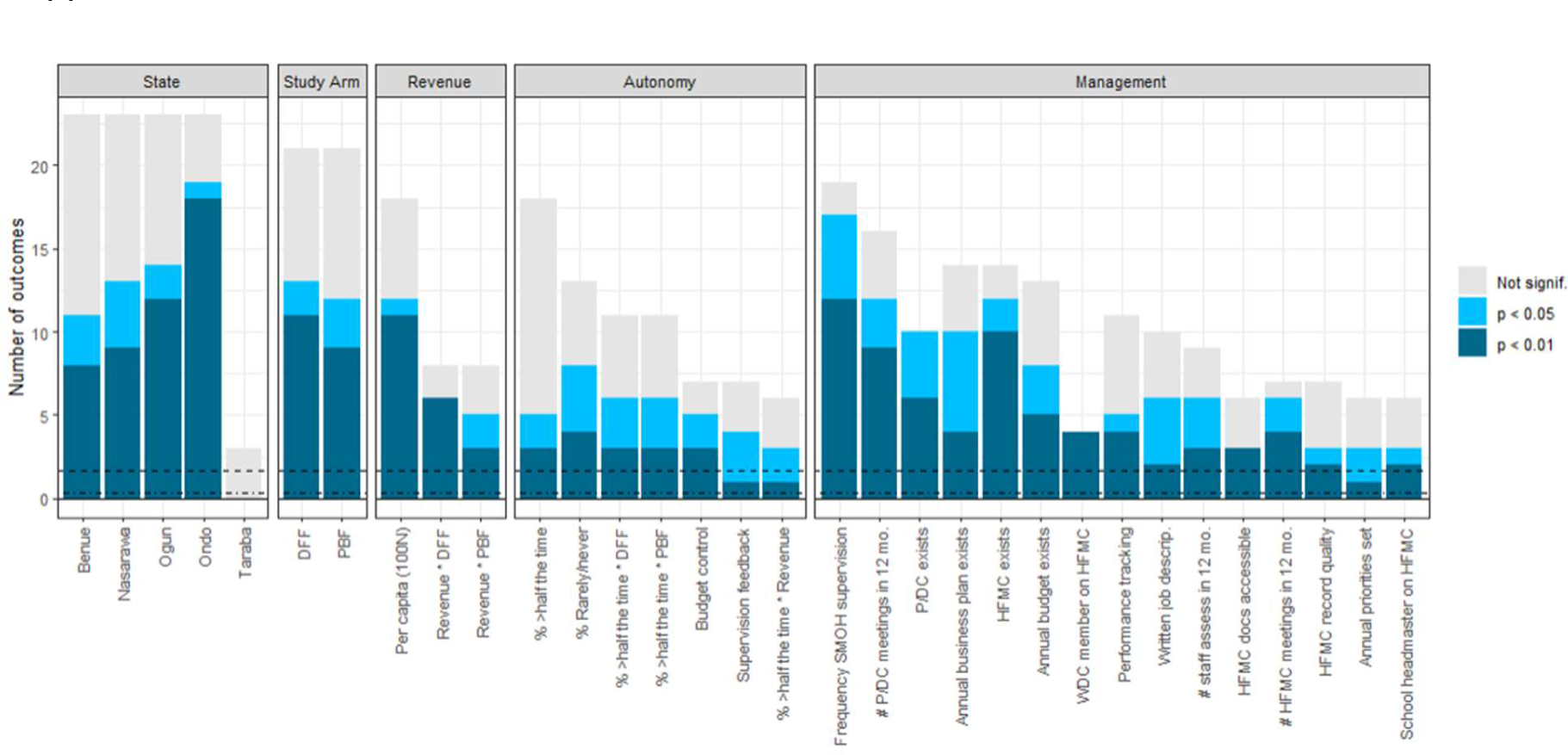
Frequency of statistical significance in predicting NSHIP facility readiness outcomes. Regressions were run using all of the variables listed on the horizontal axis as independent variables. Twenty-four outcomes were tested, which is thus the maximum possible height of a variable’s bar. Bar height represents the number of outcomes for which a variable was statistically significant at a given threshold, based on p-value. Non-significant bar indicates that a variable remained in the regression after backward selection but had a p-value higher than 0.05. Horizontal dashed lines represent the 1% and 5% levels of significance that would be expected by chance. For categorical variables, Adamawa is the baseline state and Control is the baseline study arm. DFF = direct facility financing. PBF = performance-based financing. SMOH = state ministry of health. P/DC = PBF/DFF committee. HFMC = health facility management committee. Not signif. = not significant at p<0.05 but still included in the parsimonious model after backwards variable selection.

Adjusted R-squared values ranged from 0.05 to 0.53, meaning that the regressions explained between 5% and 53% of the variation in outcomes (see Supplement 3). The more tangible items generally had higher adjusted R-squared values, such as equipment (0.49), infrastructure (0.37), and publicly posted information (0.53). For comparison, service offerings were generally positively affected by management and autonomy, but the overall power of the regressions were lower; for example, the number of outreach services offered (0.27) and frequency of vaccination outreach (0.15).

Which variables were significant predictors varied, even amongst outcomes that were related. For example, overall high autonomy was predictive of more outreach services being offered, with additional variables including P/DC meetings, annual business plan, and frequency of supervision also positively associated. High autonomy, supervisor feedback, having an annual business plan, and study arm were positively associated with holding more frequent vaccination outreach sessions. Breadth of pregnancy services (e.g. antenatal and postnatal care) and the quality of the facility’s infrastructure (e.g. walls, fence, electricity) are also shown as examples in Figure 5. Of note, the “% rarely/never” is the only variable with a negative connotation, so interpretation of directionality is reversed from the others. Overall, coefficients for management practices and revenues were mostly positive, although not all had large effect sizes. (See Supplement 3 for all coefficient estimates, significance, and uncertainty.)

**Figure 5.**
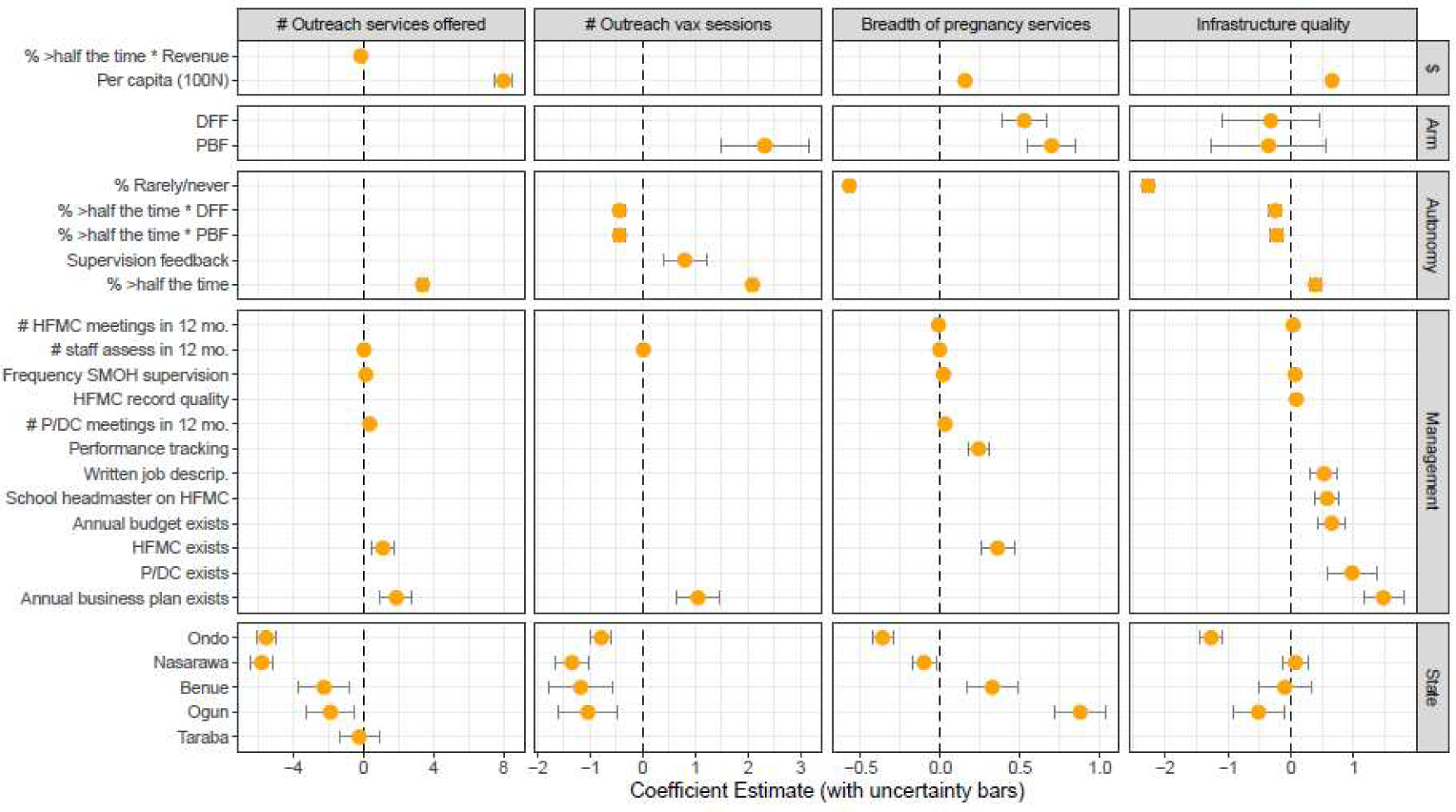
Coefficient estimates, for example regressions predicting facility readiness based on revenues, study arm, supervised autonomy and management practices. The position of the points indicates the size and direction of the effect size, only showing variables that were included in the parsimonious model. Error bars indicate the uncertainty in the estimate of the coefficient. Variables with error bars that cross the “0” line were not statistically significant. For categorical variables, Adamawa is the comparator state and Control is the default study arm. DFF = direct facility financing. PBF = performance-based financing. SMOH = state ministry of health. P/DC = PBF/DFF committee. HFMC = health facility management committee. Not signif. = not significant at p<0.05 but still included in the parsimonious model after backwards variable selection.

### Sensitivity Analysis

Regressions using the control variables only (state, study arm, revenues) performed consistently worse than those that included autonomy and management variables. All 24 of the regressions without the autonomy and management variables had higher residual sum of squares (RSS) and only two had equivalent adjusted R squared values, indicating a worse fit model without autonomy and management being included. ANOVA tests comparing the two formulations showed that these differences were statistically significant at p < 0.05 for all regressions and at p < 0.01 for 23 of 24 regressions. The adjusted R squared was meaningfully higher for some measures of readiness, such as 0.49 vs. 0.33 for general equipment, 0.41 vs. 0.22 for delivery room readiness, and 0.33 vs. 0.21 for clinical guidelines being available in the facility. (Figure 6).

**Figure 6.**
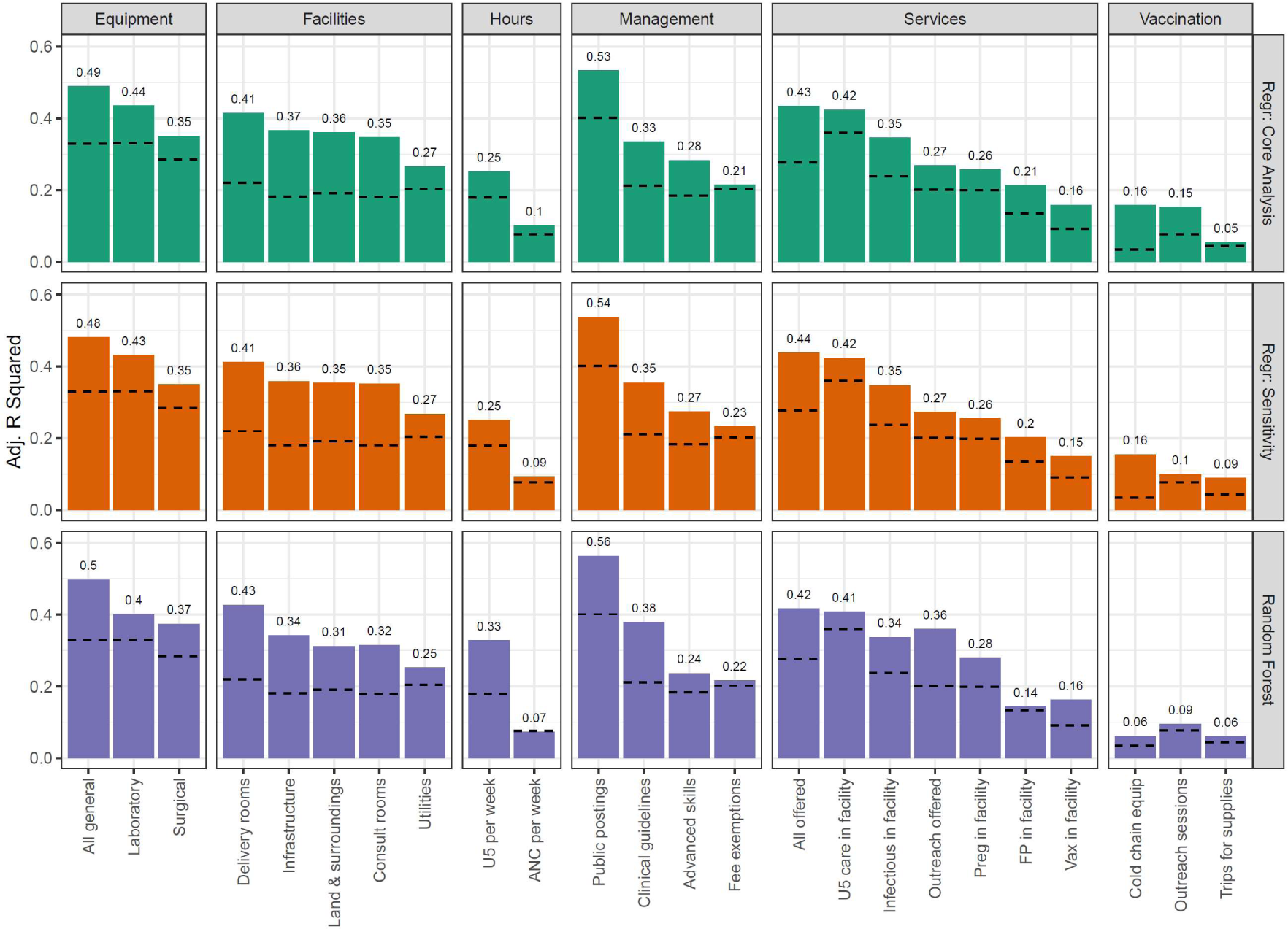
Adjusted R-squared values for sensitivity analyses, compared to core analysis results. Null model results (fit with control variables only) are shown as dashed lines for comparison to the expanded models. Core analysis regression is as described in the methods section. Sensitivity regression used the original survey values without any aggregation. Random forest model used the same variables as the core analysis but applied a different method. U5 = under 5. ANC = antenatal care. Preg = pregnancy-related. FP = family planning. In facility = the service is offered in the PHC facility. Outreach = the service is offered in the community setting. Equip = equipment.

Regressions using the unaggregated measures of autonomy and supervision performed worse on eight outcomes, equivalently on nine outcomes, and better on five outcomes. None of these were substantial differences, all within 0.05 points of each other.

The random forest model allowed for non-linearity in the model fit, but this did not substantially improve the adjusted R-squared values compared to the regression, nor did it point to structurally different results. Random forest models report “importance scores”, which estimate the contribution for independent variables *in comparison to other variables* included in the model, but they cannot be interpreted outside of that specific context. Thus, in order to use these values, we calculate the ratio of importance scores in comparison to state, which we expected to be a significant predictor given the federated nature of Nigeria’s healthcare system. Thus, we can say that on average, revenues per capita were more than twice as important as state in predicting outcomes, while study arm was 55% as important, aggregate high autonomy scores were 69% as important, aggregate low autonomy scores were 45% as important, and budget control and supervision on their own were additionally 13% as important as state.

Detailed ANOVA and random forest results can be found in Supplement 5.

Overall, the adjusted R-squared values followed similar patterns in the sensitivity analyses as in the core analysis, with the sensitivity regressions performing slightly worse and the random forest slightly better (Figure 5).

## Discussion

### Correlations

The presence of some level of correlation implies a logical relationship between certain variables (e.g. between clear and useful policies). In this analysis, we found relationships between related questions, which were to be expected. Overall, we did not find high collinearity among questions, and therefore conclude that it is acceptable to use the questions as independent variables.

### Impact of Supervision

We found that both the frequency and structure of supervision impacted facility provision of care, suggesting that routine supportive supervision has value for PHC facility performance. This happened in two ways.

First, we observed that clinical staff reported higher productivity if they had more frequent supervisory visits with the use of the quantitative checklist, especially if it exceeded a monthly cadence, which supports the findings of previous qualitative research that integrated supportive supervision can be helpful. While the magnitude of the effect was not large (∼25%), it is still a meaningful difference from a practical perspective.

Second, in DFF and PBF facilities, availability improved for all equipment items that were listed on the quarterly supervision checklist, but this was not universally true for items not on the checklist. This implies that the act of supervisors routinely reviewing equipment availability affected facility managers’ choices, to make sure that they were prepared for the supervisors’ review. This was true for both DFF and PBF facilities, even though DFF facilities were not directly incentivized for equipment availability, which suggests that the combination of supervision and funding motivated investments, so while PBF outperformed DFF, financial incentives were not the only driver of the observed improvement.

Interestingly, availability of some equipment items that were not directly incentivized also improved, reflecting that facility managers chose to spend on these items for other reasons, presumably because they were useful to support their service provision goals. The biggest improvements were seen on items supporting in-facility births, such as ringer lactate, ambubags, and the fetoscope; presumably, these choices were in response to perceived community needs for improved quality of obstetric care.

### Impact of Control over Budgets

In theory, facility managers with higher autonomy and control over how their budgets are allocated could have chosen to invest in facility readiness. We examine whether this is true for one outcome – functional equipment in the facility – and calculate this for subgroups, broken out by levels of supervised autonomy and budget control. We looked to see if increases were consistent across all facilities, or if some sub-groups of facilities had more gains than others. For facilities with at least some autonomy, and that reported high levels of control over how their budget is spent, we see substantial increases in equipment availability. However, facilities without budget control do not see the same levels of gains.

And this is even worse at lower levels of aggregate autonomy (measured across all nine questions). From this, we begin to understand that facility-level budget control was a key component of the intervention.

### Regression results

We examined the effect of autonomy and supervision on facility readiness and service offerings by running regressions on 24 aggregate measures from the survey, which covered a wide range of outcomes from waste management to stock-out rates. We found that the control variables of state, study arm, and facility revenues were highly statistically significant, which is expected since there are differences in socioeconomics and governance. The intervention overall (including incremental financing, increased autonomy, and management training) was found to be effective, and revenues can be used to solve site-specific issues, such as repairing a computer or stocking additional supplies. The interaction terms between revenue and study arm was also significant, suggesting that facility managers who were in the intervention arm of the study used their available revenues to better effect than those in the control arm.

Most of the supervised autonomy and management practice variables were significant in between a quarter and half of the regressions. All four autonomy variables were predictive, including budget control and quality of supervision. Management practices were more often predictive than autonomy. Most frequent were external supervision and the existence and functioning of the PBF/DFF and health facility management committees. Additionally, tactics such as performance tracking, having written job descriptions, and annual budgeting and business planning were also significant. The frequency is on par with (and sometimes exceeded) revenues, an expected important driver of facility functioning. This leads us to believe that autonomy and management were indeed important contributors to the outcomes seen in NSHIP, as was reported qualitatively.

It is important to note that because we are running many regressions, some variables will be significant just by chance. After removing those variables that did not exceed the 5% threshold, we can be reasonably confident that those that remain are indeed indicating a real effect.

Further, the coefficient estimates for management, supervision, and autonomy were positively associated with improved outcomes overall. We note that the management practices and revenues that formed a core part of the NSHIP intervention had positive impacts on facility outcomes. In addition, the negative relationship between facilities reporting that they “rarely or never” had autonomy and outcomes suggests that those that were disempowered had especially bad results.

There is some nuance required, however, in interpretation of these estimates, since some of these independent variables are not mutually exclusive. For example, study arm is tightly linked to level of revenues and autonomy, potentially explaining why some of the coefficients are somewhat negative for some of these variables.

By comparison, we found that autonomy was not a significant predictor of total self-reported outpatient visit volumes. This difference in results between facility readiness versus visit volumes suggests that while autonomy and good management is helpful, it is not enough on its own to result in increased service coverage. There may also be due in part to a lag between facility readiness and demand, although we cannot test this hypothesis in this study and future work is needed to examine this potential interplay.

### Sensitivity analysis

The sensitivity analysis shows that the formulation of the regression model described in the main methods section was appropriate and effective. This comes from two comparisons.

First, the addition of autonomy and supervision variables to the regression increased its prediction power. Second, the aggregation of the nine autonomy and supervision variables into summary statistics resulted in somewhat better regressions (improved 8, made 5 slightly worse) than leaving them as originally reported in the survey. This mixed effect means that, at a minimum, we conclude that the formulation we chose was not a problem for the analysis. Additionally, the random forest model had generally similar findings to the regressions, in that revenues, autonomy, management, and study arm are all important contributors to predicting facility outcomes. The random forest model does have slightly higher adjusted R-squared values, which is routinely found when comparing to a regression method, but it is not as directly interpretable.

Comparing the sensitivity analysis (i.e. without autonomy or management included) to the core regressions results, we observe that study arm is less significant and has a smaller coefficient size when autonomy and management are included. This suggests that at least part of the effect of participating in the study was due to the change in policies and practices, and not only the financial resources that were provided. Essentially, the comparison shows that study arm was less predictive once we account for supervised autonomy and management practices, so these were in fact core to the intervention and meaningfully contributed to the observed improvements in outcomes reported by the original study. This supports qualitative evidence previously reported.

### Limitations

There were several limitations to this study. Facility revenues were only documented at endline and were expected to be minimal at baseline. Causal inferences thus depend on the assumption that the randomization process resulted in similar pre-study revenues in the study arms. We believe this is likely to have been the case but cannot be sure. Secondly, many of the survey questions were asked of the officer in charge and were thus perceptions (e.g. do you receive useful feedback from your supervisor) rather than objective reports (e.g. number of immunizations administered). This introduces variability into the data and reduced our likelihood of finding an effect, so our results may be an underestimate of the true effect size. Lastly, there was the possibility that other unmeasured factors such as policy differences could have affected the facilities, but this should have been mostly mitigated by the randomization used in the original study.

### Barriers to Implementing DFF

We have demonstrated the effectiveness of DFF compared to centralized funding of PHC. Implementing DFF does however face some challenges including: (i) change management within bureaucracies who may be reluctant to ensure timely transfer of funds to front-line facilities; (ii) building capacity for financial management at PHC level and confirming compliance with public financial management rules; (iii) ensuring that all transfers of funds are electronic to facilitate auditing, record-keeping and reporting; and (iv) developing means for individual facilities to procure drugs, supplies, and equipment at prices and levels of quality that are acceptable, such as procurement framework contracts with the private sector. While these challenges are real, they have been overcome in several countries, including Tanzania and Nigeria.

### Policy Implications

The policy implications of these findings are two-fold. First, the results suggest that autonomy and supervision may have strong effects on facility readiness, so any financial reforms should consider how to incorporate some level of facility-level autonomy over planning, budgeting and decisions on the use of operational funds so that they can address readiness gaps and respond flexibly to local needs. Of note, the value of the aggregate measures suggests that a single change – such as better policy documents - is not adequate, but it should be combined with a package of changes – such as increased operational funds paired with increased supervision and operational guidance – in order to be most effective. Second, measuring management systems has been a difficult thing to do and the usefulness of these survey questions in predicting facility performance suggests that there are some aspects of PHC management that can be monitored and improved over time. Therefore, government and other stakeholders should ensure measures are put in place to monitor PHC management and use the findings to improve practice for better outcomes.

### Conclusion

Based on these analyses, we conclude that multiple aspects of supervision quality, facility autonomy, and control over budget allocation are all associated with improved facility performance. This supports qualitative findings previously reported and makes a strong case for investments in ensuring that financing reforms incorporate these enablers to support more efficient spend.

Measures like those found in this survey are not routinely collected and, given their usefulness in predicting facility outcomes, asking the facility manager about whether they receive useful supervision and other aspects of the management system may be worth incorporating into other survey tools in the future. This could become one way to track the progress of PHC systems on building their management capacity, with the goal of improving accountability. Certainly, further refinement of the measures could be useful, especially with an eye toward avoiding gaming and bias, but these existing measures can serve as a starting point for tracking PHC systems.

## Data availability

The data underlying this article are available upon request through the standard data management processes of the World Bank’s microdata library. The data ID is NGA_2017_HRBFIE-EL_v01_M and can be requested via the forms located at: https://microdata.worldbank.org/index.php/home.

## Author contributions

BH conceptualized the work, conducted data analysis and interpretation, drafted and revised the article. BL contributed to revision of the analysis, interpretation of results, and critical revisions. OO played a critical role in the data collection (during the original study), interpretation of results, and critical revision of the article. All authors have reviewed and approved the final manuscript.

## Ethical approval

No ethical approval was required for this type of study by our institutions.

## Acknowledgements

The authors would like to thank Eeshani Kandpal for providing data cleaning and access; Caitlin Mazzilli, Kevin McCarthy, and Valentina Martufi for their input into framing the initial hypotheses that motivated the analyses; and Jeremy Cooper, Rui Han, and Mandy Izzo for their editorial review.

## Supplement 1 - Supervised autonomy & management practices survey questions

**Section 2** of the Health Facility Survey. Respondent: head of the health facility or his/her deputy if absent or unavailable.

2.01 Is there a hospital/health center results based financing (PBF/DFF) committee for this health facility?

2.03 a-d Is there a representative of [health facility director / health facility staff / ward development council / headmaster of local school] on this committee?

2.04 In the last 12 months, how many hospital/health center PBF/DFF committee meetings were held?

2.05 Is there a hospital/health center management team for the facility?

2.06 In the last 12 months, how many management team meetings were held?

2.07 Are reports (business plan, minutes of meetings) of the HMIS stored in a cupboard and in a box and accessible by the administrator?

2.08 Does the facility have written records of the hospital/health center meetings?

2.09 Indicate which of the following were included in the written records? (date of meeting, agenda, list of participants, follow-up of decisions taken in the previous meeting, in each issue section there is a description of the problem, actions developed to resolve the problem, financial balance discussed, minutes of the meeting are signed)

2.11 Has a facility budget been developed for the current financial year?

2.12 Does the facility have a business plan or activity developed for the current year?

2.19 Are priority health-related activities identified for the current year?

2.22 Do all facility staff have written job descriptions?

2.23a/b/24 In the last 3 months, how many visits were made by the [SMOH/LGA/PHC department / HMB/SMOH / donor] for supervision or technical support?

2.25 In the last 12 months, how many times was the performance of staff assessed internally, that is, by persons within the facility?

2.44 Does the facility track its performance for any set of indicators?

**Section 6** of the Health Facility Survey. Text in [brackets] refers to the abbreviated label used in the results section and is provided as a cross-reference.

**Respondent**: Health facility manager/leader only

**Instructions**: *In this part of the questionnaire I would like to ask you some questions regarding how work is organized and decisions are made in this facility. All answers are confidential.*

*I am now going to read you a series of statements about decision-making and authority in this facility. Please tell me whether you feel these are true most of the time, more than half of the time, less than half of the time, rarely or never.*

6.01: I am able to allocate my facility budget according to how it is needed. There is enough flexibility in my budget. [Budget flex]

6.02: I am able to assign tasks and activities to staff as needed to achieve the outcomes I want in the facility. There is enough flexibility to use staff to address needs. [Staff assign tasks]

6.03: The HMB/LGA/PHC Department Team supports my decisions and actions for doing a better job in my facility. [HMB LGA PHC support]

6.04: I have choice over who I assign for what tasks. [Staff assign tasks]

6.05: I have choice over what services are provided in the facility. [Services offered]

6.06: I have enough authority to obtain the resources I need (drugs, supplies, funding) to meet the needs of my facility. [Obtain resources]

6.07: The policies and procedures for doing things are clear to me. [Policies clear]

6.08: The policies and procedures for doing things are useful tools for the challenges I face in providing services and reporting on activities. [Policies useful]

6.09: The HMB/LGA/PHC Department Team provides adequate feedback to me about my job and the performance of my facility. [HMB LGA PHC feedback]

## Supplement 2 – Outcome measures

**Table S2.1.**
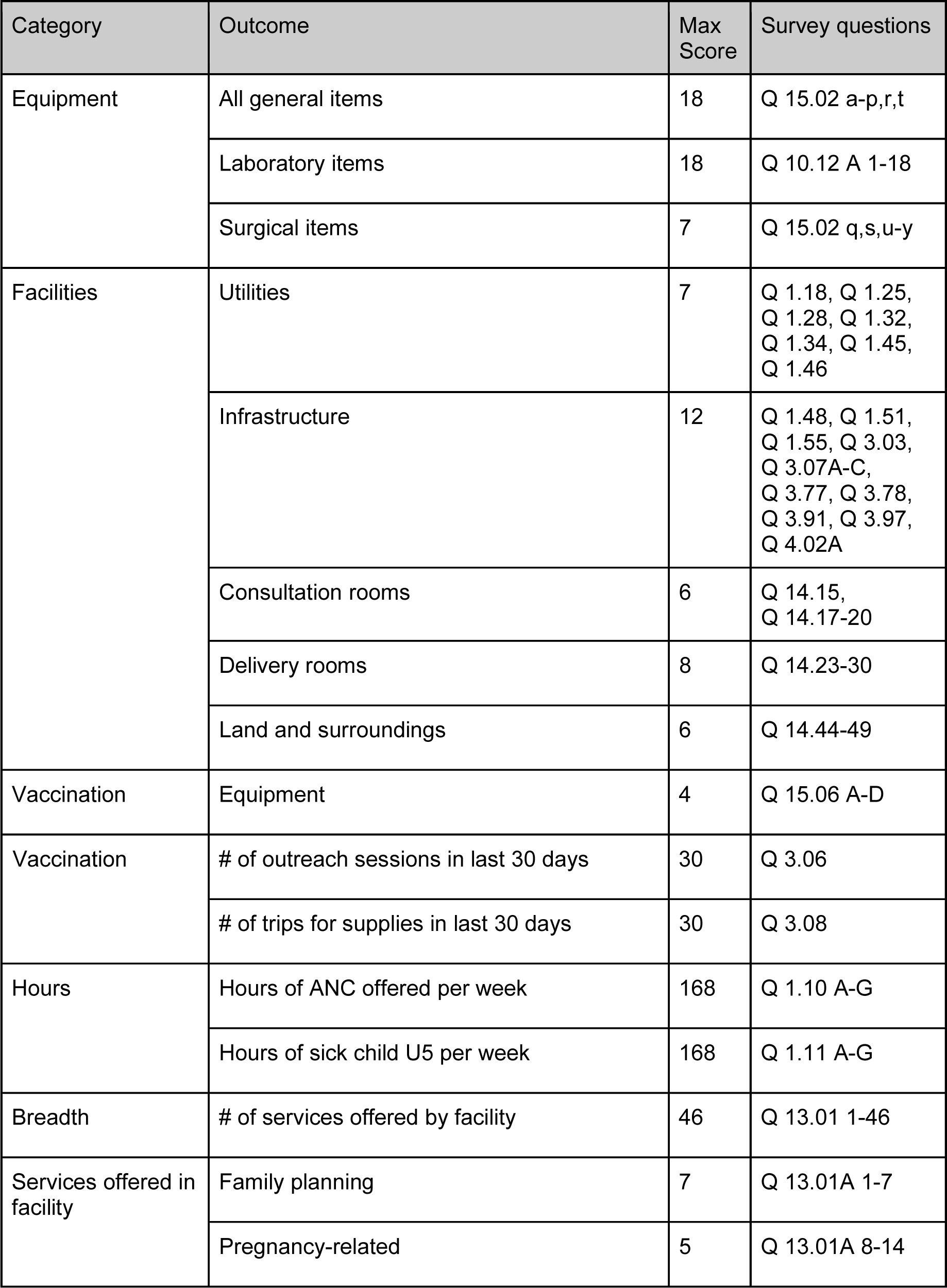
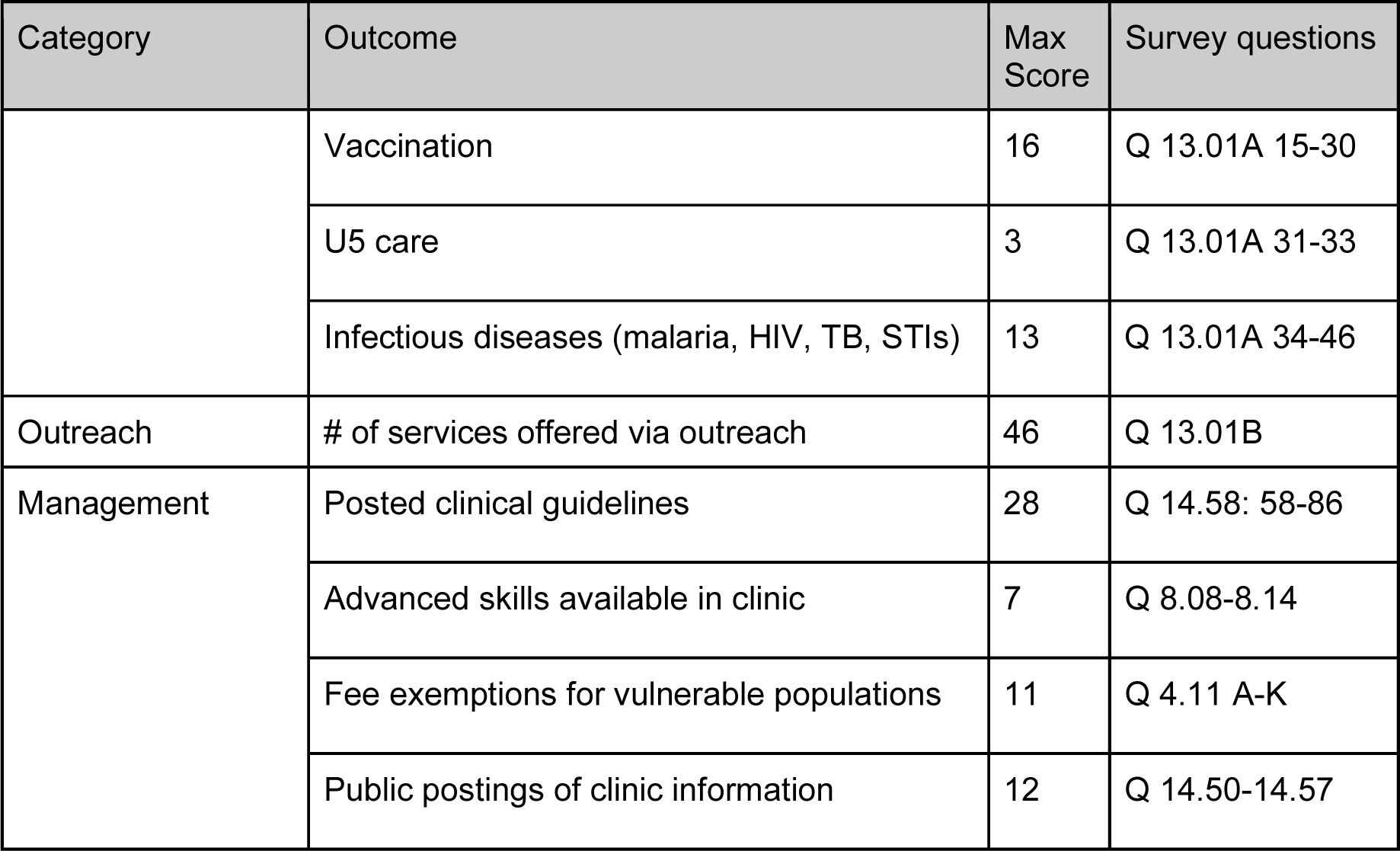
Facility outcome definitions used in regression analysis. Max score is the maximum possible facility performance score, based on survey question responses. Survey questions references the facility survey question identifier found in the endline survey documentation. ANC = antenatal care. U5 = under age 5. HIV = human immunodeficiency virus. TB = tuberculosis. STI = sexually transmitted infection. Q = question.

| Category | Outcome | Max Score | Survey questions |
| --- | --- | --- | --- |
| Equipment | All general items | 18 | Q 15.02 a-p,r,t |
|  | Laboratory items | 18 | Q 10.12 A 1-18 |
|  | Surgical items | 7 | Q 15.02 q,s,u-y |
| Facilities | Utilities | 7 | Q 1.18, Q 1.25, Q 1.28, Q 1.32, Q 1.34, Q 1.45, Q 1.46 |
|  | Infrastructure | 12 | Q 1.48, Q 1.51, Q 1.55, Q 3.03, Q 3.07A-C, Q 3.77, Q 3.78, Q 3.91, Q 3.97, Q 4.02A |
|  | Consultation rooms | 6 | Q 14.15, Q 14.17-20 |
|  | Delivery rooms | 8 | Q 14.23-30 |
|  | Land and surroundings | 6 | Q 14.44-49 |
| Vaccination | Equipment | 4 | Q 15.06 A-D |
| Vaccination | # of outreach sessions in last 30 days | 30 | Q 3.06 |
|  | # of trips for supplies in last 30 days | 30 | Q 3.08 |
| Hours | Hours of ANC offered per week | 168 | Q 1.10 A-G |
|  | Hours of sick child U5 per week | 168 | Q 1.11 A-G |
| Breadth | # of services offered by facility | 46 | Q 13.01 1-46 |
| Services offered in facility | Family planning | 7 | Q 13.01A 1-7 |
|  | Pregnancy-related | 5 | Q 13.01A 8-14 |
|  | Vaccination | 16 | Q 13.01A 15-30 |
|  | U5 care | 3 | Q 13.01A 31-33 |
|  | Infectious diseases (malaria, HIV, TB, STIs) | 13 | Q 13.01A 34-46 |
| Outreach | # of services offered via outreach | 46 | Q 13.01B |
| Management | Posted clinical guidelines | 28 | Q 14.58: 58-86 |
|  | Advanced skills available in clinic | 7 | Q 8.08-8.14 |
|  | Fee exemptions for vulnerable populations | 11 | Q 4.11 A-K |
|  | Public postings of clinic information | 12 | Q 14.50-14.57 |

## Supplement 3 - Detailed analysis results

<See Excel file.>

## Supplement 4 - Summary statistics for control vs. intervention facilities, by budget control and supervised autonomy subsets

Dataset: Number of equipment items calculated based on the potential list of 25 from the facility survey questions 15.02 A-Y. Budget control classification based on question 6.01. Autonomy classification based on questions 6.01-6.09.

**Figure S4.1.**
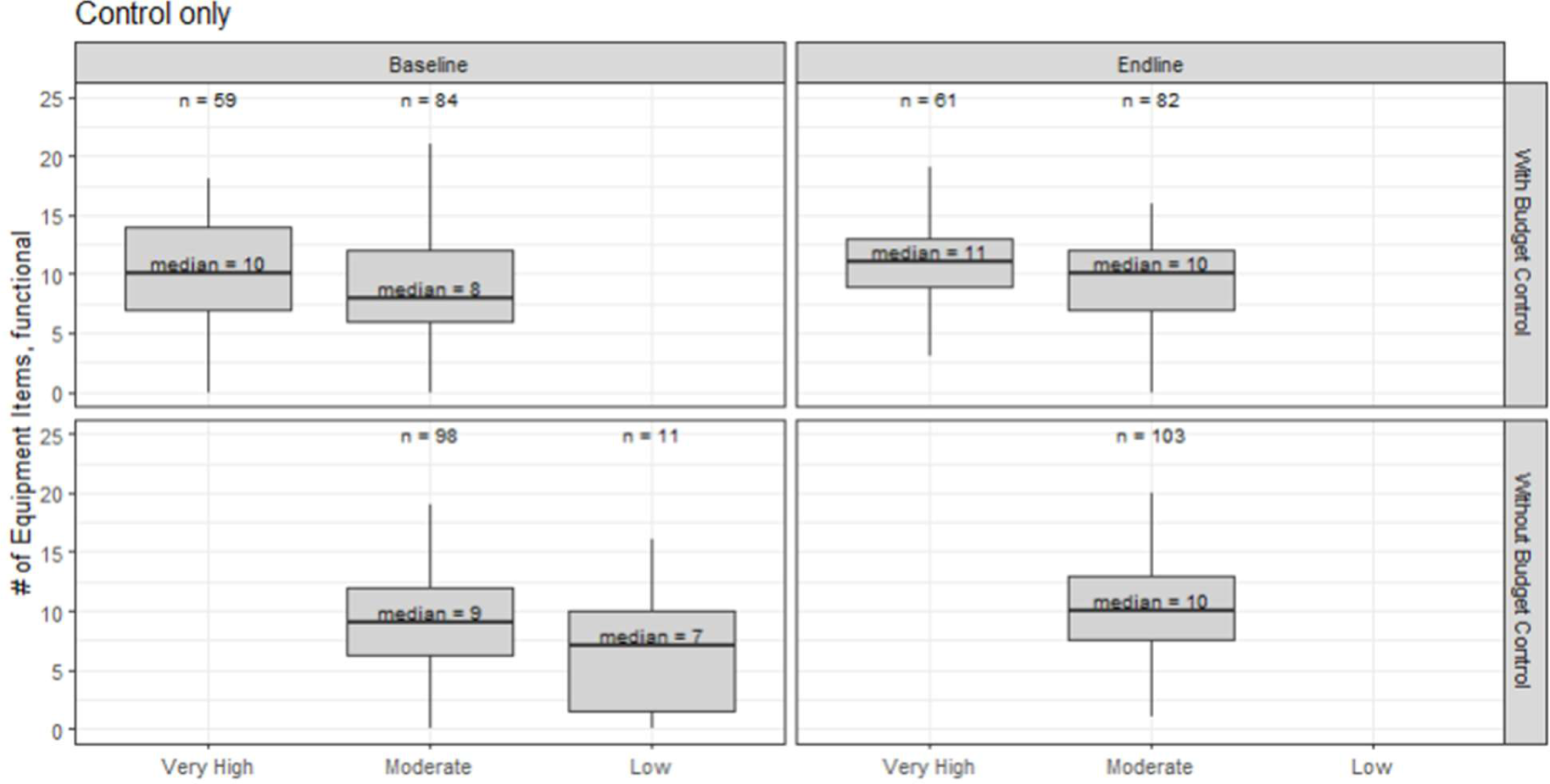
Control facilities: Impact of budget autonomy in combination with supervised autonomy on equipment availability. n = sample size. Control states: Benue, Ogun, Taraba. Boxplot central line is the distribution median, box upper and lower edges depict 25th and 75th percentiles, and lines extend to the 5th and 95th percentiles.

**Figure S4.2.**
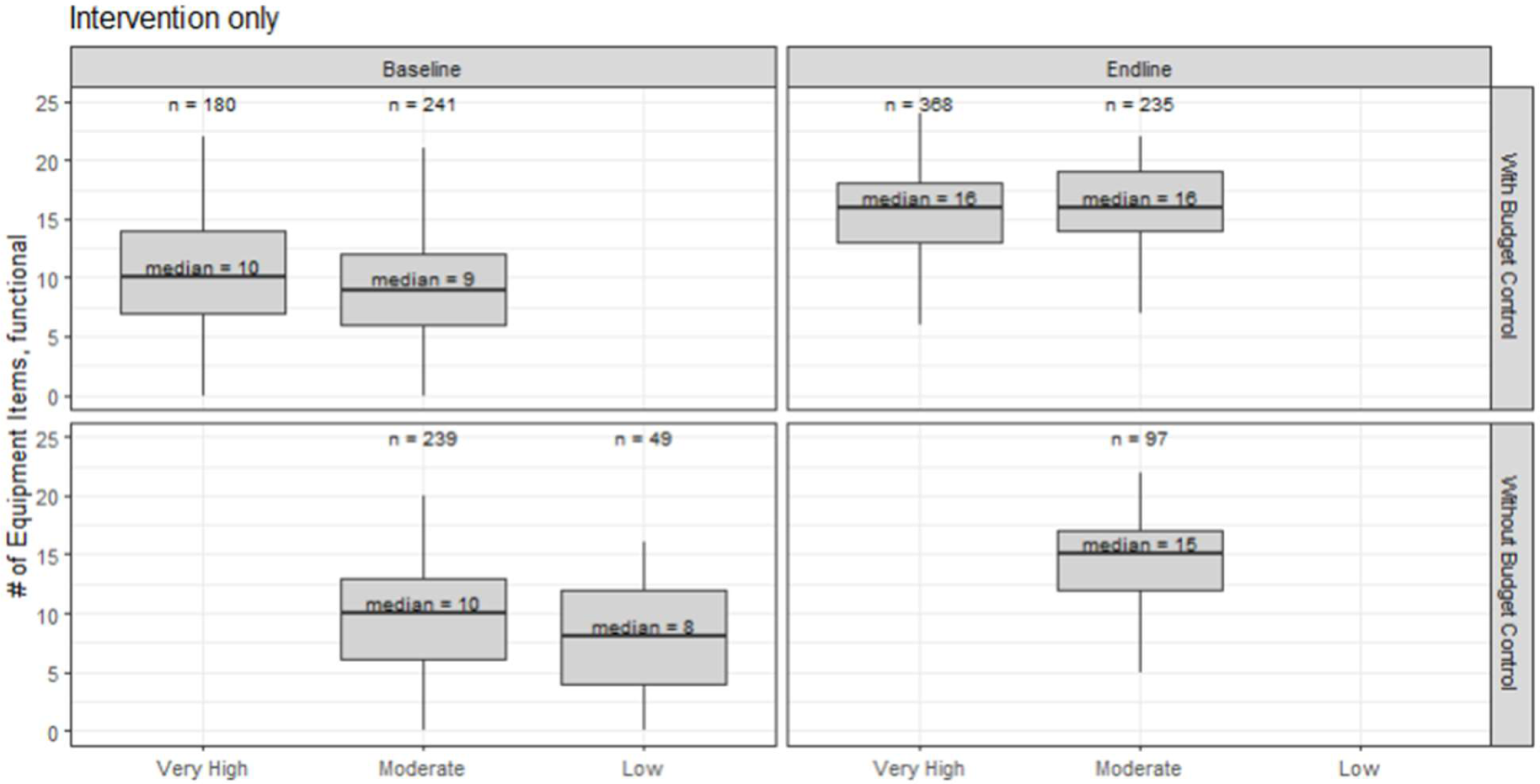
Intervention facilities: Impact of budget autonomy in combination with supervised autonomy on equipment availability. n = sample size. Control states: Adamawa, Nasarawa, Ondo. Boxplot central line is the distribution median, box upper and lower edges depict 25th and 75th percentiles, and lines extend to the 5th and 95th percentiles.

**Table S4.1.** Wilcox test for difference in means, impact of supervised autonomy and budget control on equipment availability. Bold text indicates the difference between the facility subsets being tested. W is the test statistics resulting from the Wilcox nonparametric test of difference in means. p-value indicates statistical significance. Control states: Benue, Taraba, Ogun. Intervention states: Adamawa, Nasarawa, Ondo.

| Facility subset | Subset 1 | Subset 2 | W score | p-value |
| --- | --- | --- | --- | --- |
| Control | Baseline,<br>**Very high** autonomy,<br>Budget control | Baseline,<br>**Moderate** autonomy,<br>Budget control | 2889 | 0.09 * |
| Control | Baseline,<br>**Moderate** autonomy,<br>No budget control | Baseline,<br>**Low** autonomy,<br>No budget control | 711 | 0.08 * |
| Control | Endline,<br>**Very high** autonomy,<br>Budget control | Endline,<br>**Moderate** autonomy,<br>Budget control | 3050 | 0.02 ** |
| Control | Endline,<br>**Moderate** autonomy,<br>No budget control | Endline,<br>**Low** autonomy,<br>No budget control | 1658 | 0.46 |
| Control | **Baseline**,<br>Moderate autonomy,<br>Budget control | **Endline**,<br>Moderate autonomy,<br>Budget control | 3028 | 0.18 |
| Control | **Baseline**,<br>Moderate autonomy,<br>No budget control | **Endline**,<br>Moderate autonomy,<br>No budget control | 4570 | 0.47 |
| Intervention | Baseline,<br>**Very high** autonomy,<br>Budget control | Baseline,<br>**Moderate** autonomy,<br>Budget control | 24564 | 0.02 ** |
| Intervention | Baseline,<br>**Moderate** autonomy,<br>No budget control | Baseline,<br>**Low** autonomy,<br>No budget control | 6928 | 0.04 ** |
| Intervention | Endline,<br>**Very high** autonomy,<br>Budget control | Endline,<br>**Moderate** autonomy,<br>Budget control | 38971 | 0.04 ** |
| Intervention | Endline,<br>**Moderate** autonomy,<br>No budget control | Endline,<br>**Low** autonomy,<br>No budget control | 12982 | < 0.001 *** |
| Intervention | **Baseline**,<br>Moderate autonomy,<br>Budget control | **Endline**,<br>Moderate autonomy,<br>Budget control | 10522 | < 0.001 *** |
| Intervention | **Baseline**,<br>Moderate autonomy,<br>No budget control | **Endline**,<br>Moderate autonomy,<br>No budget control | 6341 | < 0.001 *** |

## Supplement 5 - Sensitivity analysis results details

**Table S5.1.** ANOVA results from sensitivity analysis comparing a regression with control variables only (“Controls Only”) to a regression that includes those plus supervised autonomy and management practices variables, as described in the main part of the methods (“Core Analysis”). RSS = residual sum of squares.

| Category | Outcome | Controls Only |  | Core Analysis |  | ANOVA |  |
| --- | --- | --- | --- | --- | --- | --- | --- |
|  |  | RSS | Adj R2 | RSS | Adj R2 | F stat | p-value |
| Equipment | All general | 4536 | 0.36 | 3407 | 0.49 | 23.48 | <0.001 |
|  | Laboratory | 5818 | 0.34 | 4830 | 0.44 | 12.09 | <0.001 |
|  | Surgical | 1194 | 0.32 | 1074 | 0.35 | 11.43 | <0.001 |
| Facilities | Utilities | 774 | 0.21 | 705 | 0.27 | 7.77 | <0.001 |
|  | Infrastructure | 2971 | 0.2 | 2267 | 0.37 | 22.03 | <0.001 |
|  | Consult rooms | 946 | 0.2 | 743 | 0.35 | 19.41 | <0.001 |
|  | Delivery rooms | 3141 | 0.24 | 2326 | 0.41 | 24.84 | <0.001 |
|  | Land & surroundings | 1728 | 0.22 | 1347 | 0.36 | 20.07 | <0.001 |
| Vaccination | Cold chain equip | 309 | 0.05 | 261 | 0.16 | 5.52 | <0.001 |
|  | Outreach sessions | 555 | 0.12 | 501 | 0.15 | 5.83 | <0.001 |
|  | Trips for supplies | 1149 | 0.04 | 1128 | 0.05 | 2.43 | 0.0897 |
| Hours | ANC per week | 1889 | 0.09 | 1828 | 0.1 | 5.98 | <0.001 |
|  | U5 per week | 4669 | 0.21 | 4205 | 0.25 | 8.7 | <0.001 |
| Services offered | All offered | 16205 | 0.29 | 12500 | 0.43 | 21.1 | <0.001 |
|  | FP in facility | 647 | 0.15 | 576 | 0.21 | 5.74 | <0.001 |
|  | Preg in facility | 333 | 0.21 | 306 | 0.26 | 8.75 | <0.001 |
|  | Vax in facility | 1190 | 0.12 | 1083 | 0.16 | 5.76 | <0.001 |
|  | U5 care in facility | 115 | 0.38 | 103 | 0.42 | 13.64 | <0.001 |
|  | Infectious in facility | 3655 | 0.25 | 3078 | 0.35 | 9.84 | <0.001 |
|  | Outreach offered | 23752 | 0.21 | 21540 | 0.27 | 10.44 | <0.001 |
| Management | Clinical guidelines | 35510 | 0.23 | 29383 | 0.33 | 10.55 | <0.001 |
|  | Advanced skills | 511 | 0.2 | 442 | 0.28 | 9.25 | <0.001 |
|  | Fee exemptions | 4550 | 0.2 | 4454 | 0.21 | 3.86 | 0.0041 |
|  | Public postings | 4441 | 0.41 | 3387 | 0.53 | 14.64 | <0.001 |

**Table S5.2.** ANOVA results from sensitivity analysis comparing a regression as described in the main part of the methods (“Core Analysis”) with an alternative formulation for autonomy and supervision variables as described in the sensitivity analysis (“Alt Formulation”). RSS = residual sum of squares. Adj R2 = adjusted R squared value.

| Category | Outcome | Alt Formulation |  | Core Analysis |  | ANOVA |  |
| --- | --- | --- | --- | --- | --- | --- | --- |
|  |  | RSS | Adj R2 | RSS | Adj R2 | F stat | p-value |
| Equipment | All general | 3407 | 0.49 | 3407 | 0.49 | 11.38 | 8e-04 |
|  | Laboratory | 4830 | 0.44 | 4830 | 0.44 | 1.68 | 0.1242 |
|  | Surgical | 1074 | 0.35 | 1074 | 0.35 | 1.56 | 0.2123 |
| Facilities | Utilities | 705 | 0.27 | 705 | 0.27 | NA | NA |
|  | Infrastructure | 2267 | 0.37 | 2267 | 0.37 | 3.76 | 0.0107 |
|  | Consult rooms | 743 | 0.35 | 743 | 0.35 | 6.51 | 0.0109 |
|  | Delivery rooms | 2326 | 0.41 | 2326 | 0.41 | NA | NA |
|  | Land & surroundings | 1347 | 0.36 | 1347 | 0.36 | NA | NA |
| Vaccination | Cold chain equip | 261 | 0.16 | 261 | 0.16 | 1.33 | 0.2649 |
|  | Outreach sessions | 501 | 0.15 | 501 | 0.15 | 9.26 | 1e-04 |
|  | Trips for supplies | 1128 | 0.05 | 1128 | 0.05 | 4.49 | 0.0043 |
| Hours | ANC per week | 1828 | 0.1 | 1828 | 0.1 | NA | NA |
|  | U5 per week | 4205 | 0.25 | 4205 | 0.25 | 0.81 | 0.3685 |
| Services offered | All offered | 12500 | 0.43 | 12500 | 0.43 | 6.03 | 0.0143 |
|  | FP in facility | 576 | 0.21 | 576 | 0.21 | 2.77 | 0.0175 |
|  | Preg in facility | 306 | 0.26 | 306 | 0.26 | NA | NA |
|  | Vax in facility | 1083 | 0.16 | 1083 | 0.16 | 4.64 | 0.0099 |
|  | U5 care in facility | 103 | 0.42 | 103 | 0.42 | NA | NA |
|  | Infectious in facility | 3078 | 0.35 | 3078 | 0.35 | 0.37 | 0.8296 |
|  | Outreach offered | 21540 | 0.27 | 21540 | 0.27 | 5.17 | 0.0233 |
| Management | Clinical guidelines | 29383 | 0.33 | 29383 | 0.33 | 8.43 | 0 |
|  | Advanced skills | 442 | 0.28 | 442 | 0.28 | 2.22 | 0.0311 |
|  | Fee exemptions | 4454 | 0.21 | 4454 | 0.21 | 9.25 | 1e-04 |
|  | Public postings | 3387 | 0.53 | 3387 | 0.53 | NA | NA |

**Figure S5.1.**
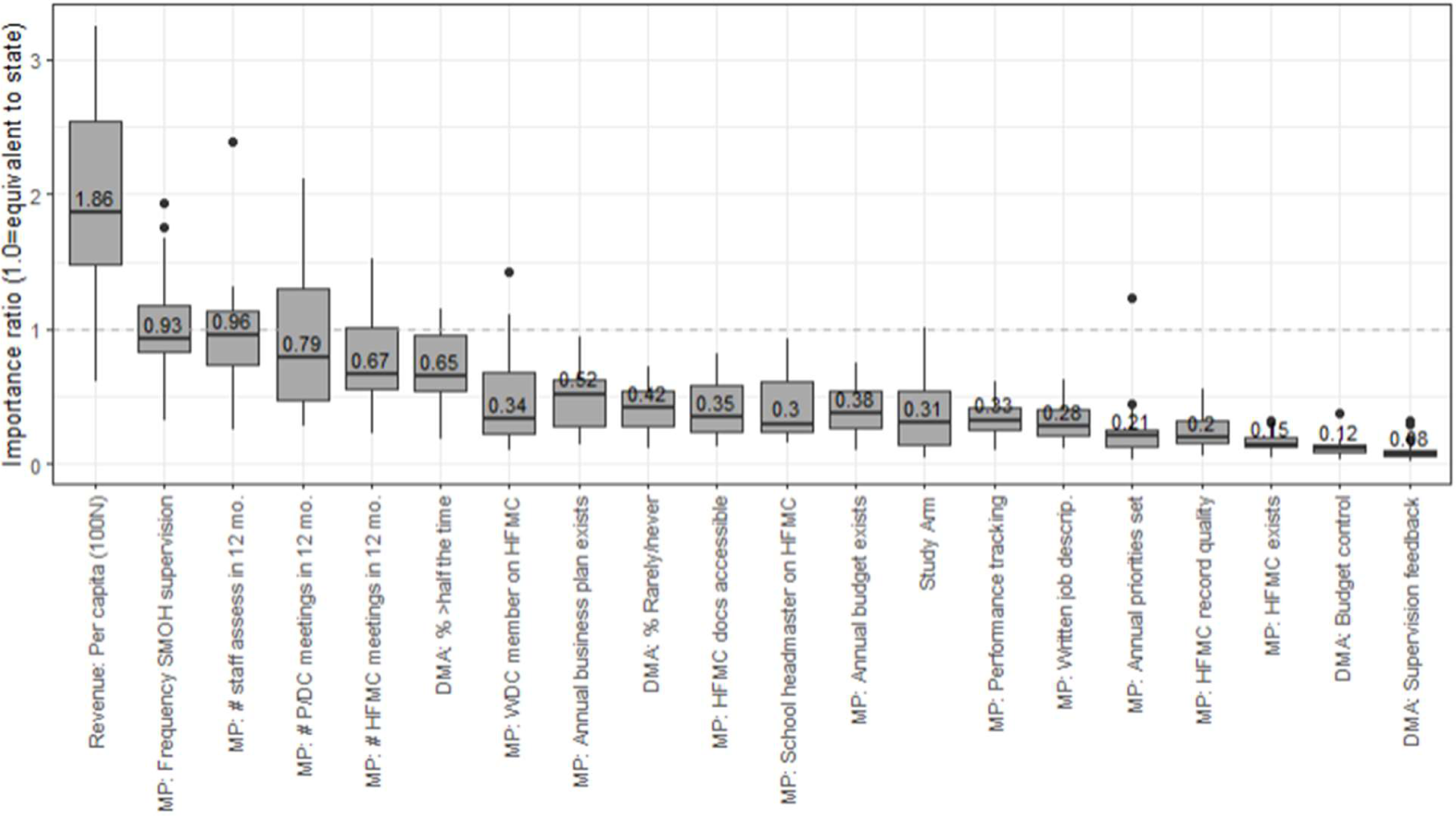
Importance scores, as a ratio to state importance, resulting from a random forest analysis using the same formulation as the core analysis in the methods section. DMA = decision making autonomy.

**Table S5.3.**
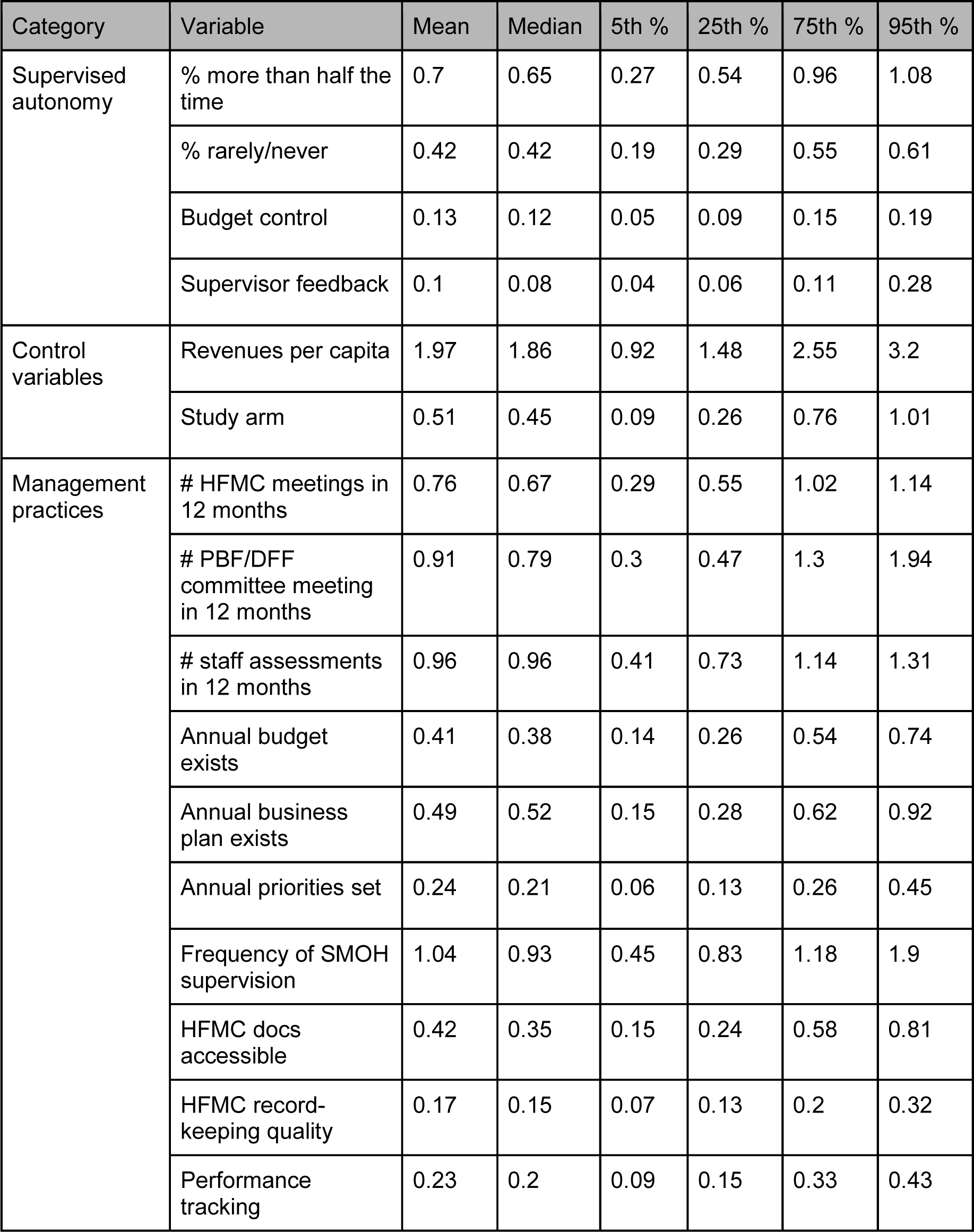
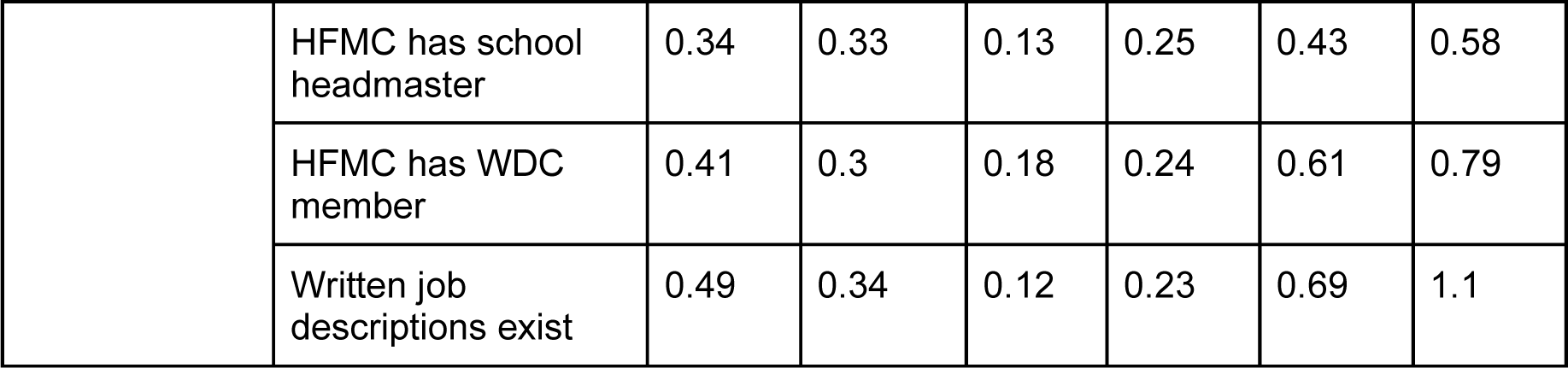
Importance scores, as a ratio to state importance, resulting from a random forest analysis using the same formulation as the core analysis in the methods section. HFMC = health facility management committee. PBF = performance-based financing. DFF = direct facility financing. SMOH = state ministry of health. WDC = ward development council. See Supplement 1 for details on the survey questions being referenced in the variables listed in this table.

| Category | Variable | Mean | Median | 5th % | 25th % | 75th % | 95th % |
| --- | --- | --- | --- | --- | --- | --- | --- |
| Supervised autonomy | % more than half the time | 0.7 | 0.65 | 0.27 | 0.54 | 0.96 | 1.08 |
|  | % rarely/never | 0.42 | 0.42 | 0.19 | 0.29 | 0.55 | 0.61 |
|  | Budget control | 0.13 | 0.12 | 0.05 | 0.09 | 0.15 | 0.19 |
|  | Supervisor feedback | 0.1 | 0.08 | 0.04 | 0.06 | 0.11 | 0.28 |
| Control variables | Revenues per capita | 1.97 | 1.86 | 0.92 | 1.48 | 2.55 | 3.2 |
|  | Study arm | 0.51 | 0.45 | 0.09 | 0.26 | 0.76 | 1.01 |
| Management practices | # HFMC meetings in 12 months | 0.76 | 0.67 | 0.29 | 0.55 | 1.02 | 1.14 |
|  | # PBF/DFF committee meeting in 12 months | 0.91 | 0.79 | 0.3 | 0.47 | 1.3 | 1.94 |
|  | # staff assessments in 12 months | 0.96 | 0.96 | 0.41 | 0.73 | 1.14 | 1.31 |
|  | Annual budget exists | 0.41 | 0.38 | 0.14 | 0.26 | 0.54 | 0.74 |
|  | Annual business plan exists | 0.49 | 0.52 | 0.15 | 0.28 | 0.62 | 0.92 |
|  | Annual priorities set | 0.24 | 0.21 | 0.06 | 0.13 | 0.26 | 0.45 |
|  | Frequency of SMOH supervision | 1.04 | 0.93 | 0.45 | 0.83 | 1.18 | 1.9 |
|  | HFMC docs accessible | 0.42 | 0.35 | 0.15 | 0.24 | 0.58 | 0.81 |
|  | HFMC record-keeping quality | 0.17 | 0.15 | 0.07 | 0.13 | 0.2 | 0.32 |
|  | Performance tracking | 0.23 | 0.2 | 0.09 | 0.15 | 0.33 | 0.43 |
|  | HFMC has school headmaster | 0.34 | 0.33 | 0.13 | 0.25 | 0.43 | 0.58 |
|  | HFMC has WDC member | 0.41 | 0.3 | 0.18 | 0.24 | 0.61 | 0.79 |
|  | Written job descriptions exist | 0.49 | 0.34 | 0.12 | 0.23 | 0.69 | 1.1 |

